# Association of caffeine consumption with memory deficits and cerebrospinal fluid biomarkers in Mild Cognitive Impairment and Alzheimer’s Disease: a BALTAZAR cohort study

**DOI:** 10.1101/2024.04.08.24305530

**Authors:** David Blum, Emeline Cailliau, Hélène Béhal, Jean-Sébastien Vidal, Constance Delaby, Luc Buée, Bernadette Allinquant, Audrey Gabelle, Stéphanie Bombois, Sylvain Lehmann, Susanna Schraen-Maschke, Olivier Hanon, the BALTAZAR study group

**Affiliations:** University of Lille, Inserm, CHU Lille, UMR-S1172 Lille Neuroscience & Cognition (LilNCog), F-59045, Lille, France; Alzheimer and Tauopathies, LabEx DISTALZ, F-59045, France; Biostatistics Department, CHU Lille, F-59045, Lille, France; Université Paris Cité, INSERM U1144, GHU APHP Centre, Hopital Broca, Memory Resource and Research Centre de Paris-Broca-Ile de France, F-75013, Paris, France; Laboratoire et Plateforme de Protéomique Clinique, Université de Montpellier, INM INSERM, IRMB CHU de Montpellier, 80 av Fliche, F-34295, Montpellier, France; Université Paris Cité, Institute of Psychiatry and Neuroscience, Inserm, UMR-S 1266, F-75014 Paris, France; Université de Montpellier, CHU Montpellier, Memory Research and Resources Center, Department of Neurology, Inserm INM NeuroPEPs Team, Excellence Center of Neurodegenerative Disorders, F-34000 Montpellier, France; Assistance Publique-Hôpitaux de Paris (AP-HP), Département de Neurologie, Centre des Maladies Cognitives et Comportementales, GH Pitié-Salpêtrière, F-75013 Paris, France; Sant Pau Memory Unit, Hospital de la Santa Creu i Sant Pau – Biomedical Research Institute Sant Pau – Universitat Autònoma de Barcelona, 08025, Barcelona, Spain

**Author notes:** Co-directed this work (David Blum, Susanna Schraen-Maschke, Olivier Hanon). Corresponding authors: Dr. David Blum, Inserm UMRS1172, Lille, France., Dr. Susanna Schraen-Mascke, Inserm UMRS1172, Lille, France.

**Keywords:** Mild cognitive impairment, Alzheimer’s disease, Caffeine, Memory, CSF biomarkers

## Abstract

**INTRODUCTION:** We investigated the link between habitual caffeine intake with memory deficits and CSF AD biomarkers in Mild cognitive impairment (MCI) and Alzheimer’s Disease (AD) patients.

**METHODS:** MCI (N=147) and AD (N=116) patients of the BALTAZAR longitudinal cohort reported their caffeine intake using a dedicated survey and were followed-up for 3 years. Associations of caffeine consumption with cognition, MMSE decline and CSF biomarkers (Tau, pTau181, Aβ_1-42_, Aβ_1-40_) were analyzed using logistic and ANCOVA models.

**RESULTS:** Adjusted on APOEε4, age, sex, education level and tobacco, lower caffeine consumption was associated with higher risk to be amnestic (OR: 2.49 [95%CI: 1.13 to 5.46]; p=0.023) and lower CSF Aβ_1-42_ (p=0.047), Aβ_1-42_/ Aβ_1-40_ (p=0.040) and Aβ_1-42_/pTau (p=0.020) in the whole cohort and a faster MMSE decline in amnestic MCI (p=0.024).

**DISCUSSION:** Data supports the beneficial effect of caffeine consumption to memory decline and CSF amyloid markers in MCI and AD patients.

## BACKGROUND

Alzheimer’s disease (AD) is characterized by a progressive cognitive decline linked to both extracellular deposits of aggregated β-amyloid (Aβ) peptides into plaques and the intraneuronal aggregation of hyperphosphorylated tau proteins [1]. Besides ageing, AD risk depends on various genetic and environmental factors [2,3].

Caffeine is the most widely consumed psychoactive agent worldwide via dietary intake from coffee, tea or soda beverages [4]. Coffee consumption has been inversely associated with total and cause-specific mortality in a large prospective cohort of participants aged 50 to 71 years at baseline (National Institutes of Health–AARP Diet and Health Study) with a 8 year follow-up of [5]. Compelling evidence supports acute caffeine’s ability to increase/improve wakefulness, alertness and memory ([6]; for review see [7,8]. Various longitudinal, cross-sectional and retrospective studies support that habitual coffee/caffeine consumption reduces cognitive decline in the elderly [9–18]; for reviews see [7] and [19]. Further, other works suggest that coffee/caffeine intake reduces dementia or AD risk [20–23]. Caffeine consumption was also associated with a decrease of behavioral symptoms in patients with dementia [24]. All these observations, mostly realized as a follow-up of non-demented elderly populations, have been acknowledged in meta-analysis studies [25–28].

Although experimental works using both amyloid and tau models showed the benefic impact of caffeine upon the development of AD lesions [29–31]; for review see [7] and [19], it remains largely unclear whether caffeine consumption associates with brain levels of amyloid and tau in MCI or AD individuals. Two studies support, in cognitively normal aged individuals a significant association between a lower coffee intake and a higher Aβ positivity as seen by PET [32,33] but these data have been discussed [34]. Notably, the link between caffeine intake and cerebrospinal fluid (CSF) biomarkers, including amyloid peptides Aβ and tau protein, in individuals presenting with MCI and AD has been largely overlooked. Only one study could not find an association between either caffeine consumption, caffeine concentration in plasma or caffeine concentration in the CSF with the levels of the core AD CSF biomarkers, in a cohort including 88 patients with AD or mild cognitive impairment (MCI) [35].

In this context, the present study aims at reinvestigating the link between habitual caffeine intake with memory deficits and CSF levels of amyloid Aβ40, Aβ42, tau and p-tau in the clinically defined BALTAZAR cohort including non-amnestic MCI (naMCI), amnestic MCI (aMCI) and AD patients.

## METHODS

### Study population

This study is ancillary to BALTAZAR (Biomarker of AmyLoid pepTide and AlZheimer’s diseAse Risk), a multicenter (23 memory centers) prospective cohort study (ClinicalTrials.gov Identifier #NCT01315639) including participants with mild cognitive impairments (MCI) and Alzheimer’s Disease (AD) at baseline from September 2010 to April 2015 and with an ongoing 3-year follow-up. All participants or their legal guardians gave written informed consent. The study was approved by the Paris Ethics Committee (CPP Ile de France IV Saint Louis Hospital). All participants were Caucasian, community-dwellers and had caregivers. Inclusion criteria for AD participants were age ≥45 years, probable AD based on the Diagnostic and Statistical Manual of Mental Disorders, 4th Edition, Text Revision and National Institute of Neurological and Communicative Disorders and Stroke and the Alzheimer’s Disease and Related Disorders Association criteria [36] and from mild to moderate stage (Mini– Mental State Examination, MMSE score ≥15). Inclusion criteria for MCI participants were age ≥70 years with MCI diagnostic criteria according to Petersen [37]. Exclusion criteria were non-AD dementia (i.e., vascular dementia, Lewy body or Parkinson dementia, and frontotemporal dementia); genetic forms of AD; major depression according to DSM IV-TR or geriatric depression scale (GDS >20/ 30); other diseases that could interfere with cognitive evaluation; diseases with short-term survival; use of cholinesterase inhibitors or methyl-D-aspartate receptor partial antagonists before the inclusion (for MCI participants); and illiterateness or less than 4 years of education.

At baseline, all the participants underwent clinical, neuropsychological, and biological assessments, and participants without contraindications underwent magnetic resonance imaging (MRI) brain examinations. Cognitive evaluations were performed by neuropsychologists after training program to harmonize the quotation. MCI and AD participants underwent every six months for three years a neuropsychological test battery that included global cognitive assessment with the Mini–Mental State Examination (MMSE, normal score 30/30), Clinical Dementia Rating Scale (CDR, CDR sum of boxes: normal score 0/18). Functional disability was assessed using instrumental activities of daily living (IADL, normal score 14/14) (for the description of all tests performed in the cohort see [38]. MCI participants were then categorized as amnestic (aMCI) or non-amnestic (naMCI) according to the presence of memory impairment on the free and cued selective recall reminding test related to age, sex, and educational level [39]. Conversion to dementia in the naMCI group was very low (2.5% i.e 1/40) as compared to the aMCI group (22.4% i.e. 24/107).

### CSF biomarker measurements

CSF biomarker levels of Tau, pTau181, Aβ40 and Aβ42, were determined with standardized commercially available ELISA Kits (Euroimmun β-amyloid 1–40 and 1–42, Innotest hTau and Innotest Phospho-Tau181). Routine CSF analyses included glucose, total protein and cytology. Only samples with red cell count <2000/mm^3^ were analysed.

### Habitual caffeine consumption

The present study enrolled 263 patients in which the habitual caffeine intake was assessed using an in-housed validated self-survey [40] filled by the patients and caregiver. The daily intake of caffeine containing items (coffee, tea, chocolate, sodas) was reported, along with any changes of consumption over the last 6 months period. Daily caffeine intake was then calculated at inclusion.

### Statistical analysis

Categorical variables are reported as frequency (percentage). Continuous variables are reported as mean (standard deviation, SD) in the case of normal distribution or median (interquartile range, IQR) otherwise. Normality of distributions was assessed using histograms and using the Shapiro-Wilk test.

Patients characteristics and CSF biomarkers were compared between the three diagnostic groups (naMCI, aMCI and DA) using Chi-square test (or Fisher’s exact test in case of expected value <5) for categorical variables and using Analysis of variance or Kruskal-Wallis test for quantitative variables.

Impact of low or high caffeine consumption on the diagnostic group was evaluated using a multinomial logistic regression model adjusted on predefined confounding factors (APOEε4, age, sex, education level and tobacco consumption). Odds ratios were derived from models with their 95% confidence intervals. The same analysis was performed by regrouping aMCI and DA groups, using a logistic regression model adjusted on predefined confounding factors.

Impact of low or high caffeine consumption on CSF biomarkers was evaluated using an analysis of covariance (ANCOVA) adjusted on predefined confounding factors and after having applied a log transformation of the CSF biomarkers. Mean difference was derived from the model with their 95% confidence intervals.

The variation of the MMSE score between baseline and 36 months was compared between the two caffeine consumption groups (low versus high) using an ANCOVA adjusted on predefined confounding factors and on baseline MMSE. Mean difference was derived from the model with their 95% confidence intervals. Due to the limited number of subjects in naMCI group, this analysis was performed only in aMCI and DA diagnostic groups.

All statistical tests were done at the two-tailed α-level of 0.05 using the SAS software version 9.4 (SAS Institute, Cary, NC).

## RESULTS

### Demographic and clinical characteristics at baseline

263 participants of the BALTAZAR cohort with available caffeine survey were included (40 naMCI, 107 aMCI and 116 AD; **Table 1**). At baseline, among the 263 participants, 38.0% (N=100) were males, 40.8% (N=106) had at least high school diploma and 74.8% (N=193) never smoked. The median MMSE score was 26 (interquartile range (IQR): 23 to 28) and 47.7% (N=114) were APOEε4 carriers. The median caffeine consumption was 216 (IQR: 84 to 374) mg/day. During the clinical follow-up period (6-36 months), 22.4% (N=24) of the aMCI and 2.5% (N=1) of the naMCI participants developed dementia and converted to probable AD. During this 3-year follow-up, 36.9% (N=179) of participants were lost to follow-up: 8.4 % (N=41) after the baseline visit, 5.8% (N=28) after the 6-month visit, 3.3% (N=16) after the 12-month visit, 3.3% (N=16) after the 18-month visit, 15.1% (N=73) after the 24-month visit, 1.0% (N=5) after the 30-month visit. Moreover, 15 participants died during the 3-year follow-up. Analysis of CSF biomarkers at inclusion showed higher CSF total Tau (Tau) and phospho-Tau (pTau) levels and lower amyloid Aβ42 levels, Aβ42/Aβ40 and Aβ42/pTau ratios in AD patients as compared to naMCI and aMCI individuals. Similarly, hippocampal volume, MMSE and IADL were significantly lower while CDR was higher in AD versus aMCI and naMCI (**Table 1**).

**Table 1:**
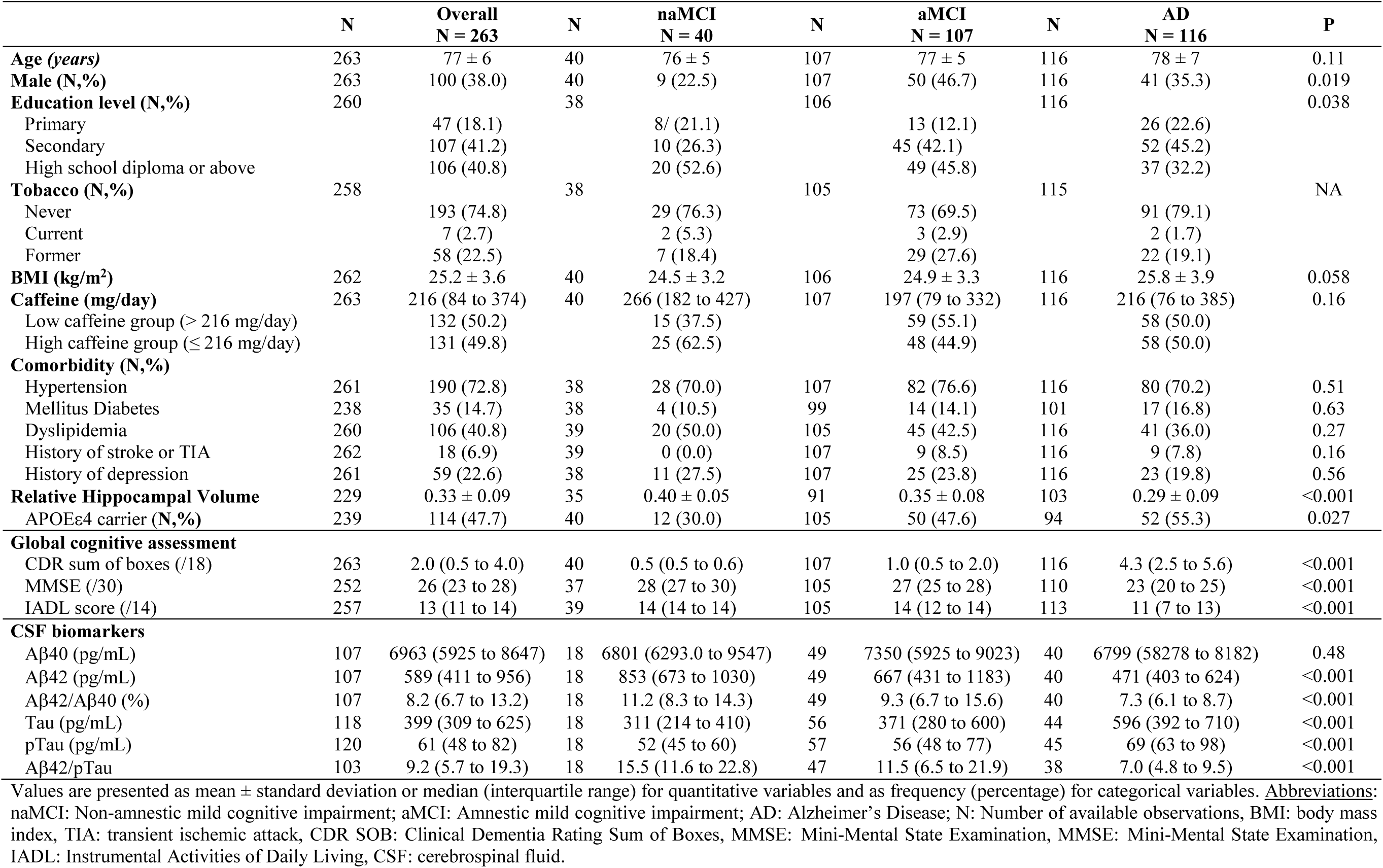
General characteristics, global cognitive assessment and CSF biomarkers at baseline.

|  | N | Overall<br>N = 263 | N | naMCI<br>N = 40 | N | aMCI<br>N = 107 | N | AD<br>N = 116 | P |
| --- | --- | --- | --- | --- | --- | --- | --- | --- | --- |
| **Age (years)** | 263 | 77 ± 6 | 40 | 76 ± 5 | 107 | 77 ± 5 | 116 | 78 ± 7 | 0.11 |
| **Male (N,%)** | 263 | 100 (38.0) | 40 | 9 (22.5) | 107 | 50 (46.7) | 116 | 41 (35.3) | 0.019 |
| **Education level (N,%)** | 260 |  | 38 |  | 106 |  | 116 |  | 0.038 |
| Primary |  | 47 (18.1) |  | 8/ (21.1) |  | 13 (12.1) |  | 26 (22.6) |  |
| Secondary |  | 107 (41.2) |  | 10 (26.3) |  | 45 (42.1) |  | 52 (45.2) |  |
| High school diploma or above |  | 106 (40.8) |  | 20 (52.6) |  | 49 (45.8) |  | 37 (32.2) |  |
| **Tobacco (N,%)** | 258 |  | 38 |  | 105 |  | 115 |  | NA |
| Never |  | 193 (74.8) |  | 29 (76.3) |  | 73 (69.5) |  | 91 (79.1) |  |
| Current |  | 7 (2.7) |  | 2 (5.3) |  | 3 (2.9) |  | 2 (1.7) |  |
| Former |  | 58 (22.5) |  | 7 (18.4) |  | 29 (27.6) |  | 22 (19.1) |  |
| **BMI (kg/m<sup>2</sup>)** | 262 | 25.2 ± 3.6 | 40 | 24.5 ± 3.2 | 106 | 24.9 ± 3.3 | 116 | 25.8 ± 3.9 | 0.058 |
| **Caffeine (mg/day)** | 263 | 216 (84 to 374) | 40 | 266 (182 to 427) | 107 | 197 (79 to 332) | 116 | 216 (76 to 385) | 0.16 |
| Low caffeine group (> 216 mg/day) |  | 132 (50.2) |  | 15 (37.5) |  | 59 (55.1) |  | 58 (50.0) |  |
| High caffeine group (≤ 216 mg/day) |  | 131 (49.8) |  | 25 (62.5) |  | 48 (44.9) |  | 58 (50.0) |  |
| **Comorbidity (N,%)** |  |  |  |  |  |  |  |  |  |
| Hypertension | 261 | 190 (72.8) | 38 | 28 (70.0) | 107 | 82 (76.6) | 116 | 80 (70.2) | 0.51 |
| Mellitus Diabetes | 238 | 35 (14.7) | 38 | 4 (10.5) | 99 | 14 (14.1) | 101 | 17 (16.8) | 0.63 |
| Dyslipidemia | 260 | 106 (40.8) | 39 | 20 (50.0) | 105 | 45 (42.5) | 116 | 41 (36.0) | 0.27 |
| History of stroke or TIA | 262 | 18 (6.9) | 39 | 0 (0.0) | 107 | 9 (8.5) | 116 | 9 (7.8) | 0.16 |
| History of depression | 261 | 59 (22.6) | 38 | 11 (27.5) | 107 | 25 (23.8) | 116 | 23 (19.8) | 0.56 |
| **Relative Hippocampal Volume** | 229 | 0.33 ± 0.09 | 35 | 0.40 ± 0.05 | 91 | 0.35 ± 0.08 | 103 | 0.29 ± 0.09 | <0.001 |
| **APOEε4 carrier (N,%)** | 239 | 114 (47.7) | 40 | 12 (30.0) | 105 | 50 (47.6) | 94 | 52 (55.3) | 0.027 |
| **Global cognitive assessment** |  |  |  |  |  |  |  |  |  |
| CDR sum of boxes (/18) | 263 | 2.0 (0.5 to 4.0) | 40 | 0.5 (0.5 to 0.6) | 107 | 1.0 (0.5 to 2.0) | 116 | 4.3 (2.5 to 5.6) | <0.001 |
| MMSE (/30) | 252 | 26 (23 to 28) | 37 | 28 (27 to 30) | 105 | 27 (25 to 28) | 110 | 23 (20 to 25) | <0.001 |
| IADL score (/14) | 257 | 13 (11 to 14) | 39 | 14 (14 to 14) | 105 | 14 (12 to 14) | 113 | 11 (7 to 13) | <0.001 |
| **CSF biomarkers** |  |  |  |  |  |  |  |  |  |
| Aβ40 (pg/mL) | 107 | 6963 (5925 to 8647) | 18 | 6801 (6293.0 to 9547) | 49 | 7350 (5925 to 9023) | 40 | 6799 (58278 to 8182) | 0.48 |
| Aβ42 (pg/mL) | 107 | 589 (411 to 956) | 18 | 853 (673 to 1030) | 49 | 667 (431 to 1183) | 40 | 471 (403 to 624) | <0.001 |
| Aβ42/Aβ40 (%) | 107 | 8.2 (6.7 to 13.2) | 18 | 11.2 (8.3 to 14.3) | 49 | 9.3 (6.7 to 15.6) | 40 | 7.3 (6.1 to 8.7) | <0.001 |
| Tau (pg/mL) | 118 | 399 (309 to 625) | 18 | 311 (214 to 410) | 56 | 371 (280 to 600) | 44 | 596 (392 to 710) | <0.001 |
| pTau (pg/mL) | 120 | 61 (48 to 82) | 18 | 52 (45 to 60) | 57 | 56 (48 to 77) | 45 | 69 (63 to 98) | <0.001 |
| Aβ42/pTau | 103 | 9.2 (5.7 to 19.3) | 18 | 15.5 (11.6 to 22.8) | 47 | 11.5 (6.5 to 21.9) | 38 | 7.0 (4.8 to 9.5) | <0.001 |

Values are presented as mean ± standard deviation or median (interquartile range) for quantitative variables and as frequency (percentage) for categorical variables. **Abbreviations:** naMCI: Non-amnesic mild cognitive impairment; aMCI: Amnesic mild cognitive impairment; AD: Alzheimer's Disease; N: Number of available observations, BMI: body mass index, TIA: transient ischemic attack, CDR SOB: Clinical Dementia Rating Sum of Boxes, MMSE: Mini-Mental State Examination, MMSE: Mini-Mental State Examination, IADL: Instrumental Activities of Daily Living, CSF: cerebrospinal fluid.

### Association of caffeine consumption with the diagnostic groups at baseline according to the cognitive status (AD, aMCI, naMCI)

Participants were dichotomized according to their median caffeine consumption (216 mg/day) in the overall cohort as well as in each subgroup (AD, aMCI, naMCI). After adjustment on APOEε4, age, sex, education level and tobacco consumption, and using naMCI group as reference, we found a significative association of a lower caffeine consumption with a higher risk to be aMCI (Odds Ratio (OR): 2.72 [95% Confidence Interval (CI): 1.17 to 6.30]) and a similar effect size, even non-significant, for higher risk of being AD (OR:2.31 [95%CI: 0.98 to 5.40]; **Table 2**). When aMCI and AD groups were combined, the association of a lower caffeine consumption with higher risk to be amnestic was significant (OR: 2.49 [95%CI: 1.13 to 5.46]; p=0.023) (**Table 2**).

**Table 2:** Association of caffeine consumption according to cognitive status (naMCI, aMCI, AD)

| **Overall** | **N = 263** | **Low caffeine consumption**<br>**N = 132** | **High caffeine consumption**<br>**N = 131** | **Odds Ratio (95% CI)** | **P** |
| --- | --- | --- | --- | --- | --- |
| **naMCI_aMCI_AD (N, %)** |  |  |  |  | 0.062 |
| naMCI | 40 (15.2) | 15 (11.4) | 25 (19.1) | 1.00 (réf.) | - |
| aMCI | 107 (40.7) | 59 (44.7) | 48 (36.6) | 2.72 (1.17 to 6.30) | **0.019** |
| AD | 116 (44.1) | 58 (43.9) | 58 (44.3) | 2.31 (0.98 to 5.40) | 0.054 |
| **naMCI_aMCI+AD (N, %)** |  |  |  |  |  |
| naMCI | 40 (15.2) | 15 (11.4) | 25 (19.1) | 1.00 (réf.) | - |
| aMCI+AD | 223 (84.8) | 117 (88.6) | 106 (80.9) | 2.49 (1.13 to 5.46) | **0.023** |

Values are presented as frequency (percentage). Odds ratios and Pvalues were adjusted for APOEε4, age, sex, education level and tobacco consumption. Abbreviations: naMCI: Non-amnestic mild cognitive impairment; aMCI: Amnestic mild cognitive impairment; AD: Alzheimer's Disease; N: Number of available observations; CI: Confidence Interval; P: Pvalue.

### Association of caffeine consumption with CSF amyloid and tau biomarkers at baseline

Among the whole cohort, after adjustment on APOEε4, age, sex, education level and tobacco consumption, lower caffeine consumption was found associated with lower CSF amyloid Aβ42 levels (p=0.047) as well as lower Aβ42/Aβ40 (p=0.040) and Aβ42/pTau ratio (p=0.020; **Figures 1A-D**). No significant difference was observed regarding CSF total Tau and pTau levels according to the caffeine consumption (**Figure 1A**).

**Figure 1.**
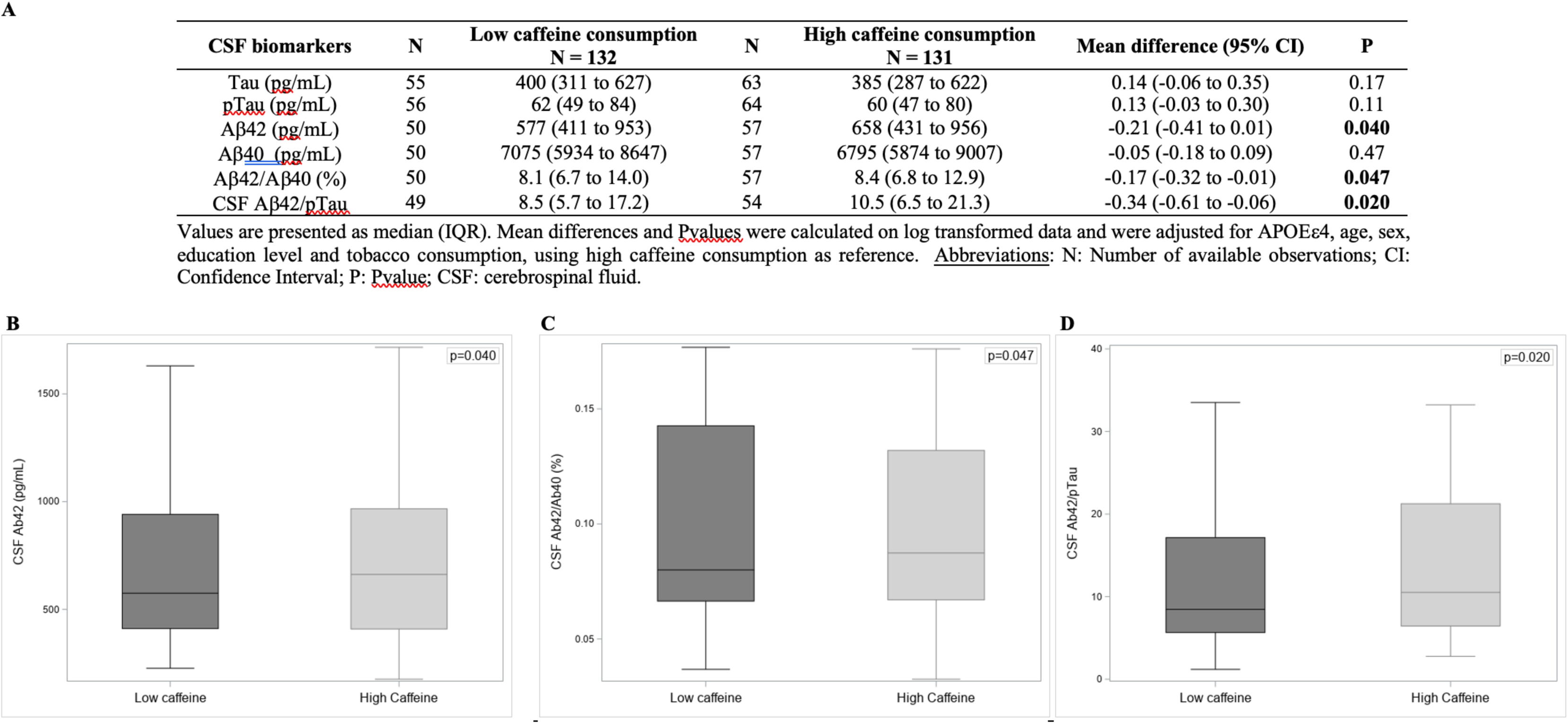
Association of caffeine consumption with CSF biomarkers at baseline. (**A**) Association between caffeine consumption and CSF Biomarkers. Values are presented as median (IQR). Mean differences and Pvalues were calculated on log transformed data and were adjusted for APOEε4, age, sex, education level and tobacco consumption, using high caffeine consumption as reference. Abbreviations: N: Number of available observations; CI: Confidence Interval; P: Pvalue; CSF: cerebrospinal fluid. **(B)** CSF Aβ1-42. **(C)** CSF Aβ1-42/Aβ1-40. **(D)** CSF Aβ1-42/pTau.

### Association of caffeine consumption with MMSE decline

We further tested whether caffeine consumption was associated with cognitive impairment measured by the decline in MMSE from baseline to 36 months (mean follow-up: 25 months; **Table 3**). After adjustment on APOEε4, age, sex, education level and tobacco consumption, the decline of MMSE was more important in low caffeine consumers than in high caffeine consumers effect size in the aMCI group (adjusted mean difference: –1.3 [95%CI: –2.4 to –0.2]; p=0.024; **Table 3**). However, the MMSE decline was not significantly different in AD subjects.

**Table 3:** Association of caffeine consumption with MMSE decline.

|  | N | aMCI<br>N = 107 | N | Low caffeine<br>consumption<br>N = 53 | N | High caffeine<br>consumption<br>N = 54 | Mean difference<br>(95% CI) | P |
| --- | --- | --- | --- | --- | --- | --- | --- | --- |
| **Slope MMSE** | 66 | -0.4 ± 2.2 | 38 | -0.7 ± 2.3 | 28 | -0.1 ± 2.0 | -1.3 (-2.4 to -0.2) | **0.024** |
|  |  | AD<br>N = 116 |  | Low caffeine<br>consumption<br>N = 57 |  | High caffeine<br>consumption<br>N = 58 | Mean difference<br>(95% CI) | P |
| **Slope MMSE** | 81 | -3.4 ± 3.8 | 41 | -3.1 ± 3.7 | 40 | -3.8 ± 3.9 | 0.3 (-1.5 to 2.0) | 0.78 |

Values are presented as mean ± SD. Mean differences and Pvalues were adjusted for APOEε4, age, sex, education level and tobacco consumption, using high caffeine consumption as reference. Abbreviations: aMCI: Amnesic mild cognitive impairment; AD: Alzheimer's Disease; SD: Standard Deviation; CI: Confidence Interval; P: P value, MMSE: Mini-Mental State Examination.

## DISCUSSION

In the present study, we evaluated the impact of habitual caffeine consumption on clinical outcomes and biomarkers at baseline as well as on memory decline in the BALTAZAR cohort including MCI and AD patients. Our data show an association of a lower caffeine consumption with memory disorders related to AD and aMCI at inclusion but also with a higher risk of MMSE decline in the amnestic MCI patients. Importantly, our data also demonstrate that caffeine associates with changes in CSF biomarkers of AD patients, particularly with regards of the amyloid component.

The association fits well with few, and potentially underpowered, previous retrospective studies investigating the impact of caffeine consumption on populations of patients presenting with MCI or AD. [21] showed a significantly lower average consumption of caffeine (74 mg/day, N=54) during the 20 years preceding AD development subjects as compared with age-matched non-demented subjects (approximately 200 mg/day, N=54). In the same way, [41] suggested that plasma caffeine concentrations were approximately 50% lower in subjects converting to dementia (N=15) (presumably comparable to our aMCI group at high risk of converting to dementia) vs. subjects that do not convert (N=9) (presumably comparable to our naMCI group). Interestingly, one study [42], in line with experimental data [43], found that higher caffeine intake was associated with better function in overall cognition, encompassing episodic memory, executive function, semantic categorization, and working memory, in a cohort of approximately 600 subjects presenting with diabetes, known to be a significant risk factor for dementia [44], as are aMCI individuals [45]. These data support a benefit of habitual caffeine consumption in at risk individuals that would warrant to extend more largely cognitive endophenotypes at MCI inclusion and follow-up per caffeine consumption. Besides the association of caffeine with memory at inclusion, we also found a significant association of lower caffeine consumption with a faster decline in MMSE in aMCI subjects but not in AD patients. To the best of our knowledge, this is the only study carried out specifically on MCI or AD subjects. However, these observations are in agreement with previous longitudinal and cross-sectional studies, including initially cognitively unimpaired population, that associate caffeine consumption to a reduced age-related cognitive decline (for reviews see [19,46]). Notably, two studies on large cohorts of non-demented elderly subjects have particularly shown a significant association of higher caffeine consumption with a lower decline in MMSE over 8-10 years [10,12]. Interestingly, our association between reduced MMSE decline and higher caffeine consumption is only observed for the aMCI individuals but not the AD patients. This suggests that the beneficial effect of caffeine on cognitive decline might remain limited to the earlier stages of pathology, presumably before pathophysiological mechanisms reach an irreversible turning point. Besides an impact on brain lesions (see below), improved cognition we observed might primarily relate to the ability of caffeine to act on synaptic plasticity. Indeed, in rodents, caffeine treatment on hippocampal slices enhances basal synaptic transmission, long-term potentiation (LTP) and sharp wave–ripple complexes, which underlie memory consolidation [47–50]. Caffeine also controls neuronal excitability and LTP-like effects in the human cortex [51,52]. More recently, we also demonstrated that regular caffeine intake act on epigenomic and transcriptional processes that would improve the signal-to-noise ratio during information encoding, favoring the function of circuits involved in learning [53]. The beneficial effect of caffeine on synaptic plasticity during MCI and AD would be particularly ascribed to its ability to block adenosine receptors [4,7] and [7] for reviews, particularly the A_2A_R subtypes [54].

Importantly, we show a significant association across the cohort between lower caffeine intake and lower CSF levels Aβ42 as well as Aβ42/Aβ40 and Aβ42/pTau ratios. To our knowledge, this is the first study reporting an association between caffeine and CSF AD biomarkers. Only one previous study unsuccessfully addressed such association in another population (Travassos et al., 2015). However, this study included a more limited number of subjects (37 MCI and 51 AD), the biomarker assay techniques were different and, for an unclear reason, the median caffeine consumption was 2 times lower (around 100 mg/day) than in our population. Using our in-housed survey – developed to get more precise consumption habit than caffeine unit or mugs – and considering all types of caffeine consumption and volume ingested, we estimated the median consumption of our population to be 216 mg/day. This amount was in line with previous studies in cohorts of a similar age [18,32,42]. Our CSF data point towards a particular impact of caffeine on amyloid burden. This is in accordance with a Korean study [33] including elderly subjects (mean age 71 years-old) without dementia (282 cognitively normal and 129 MCI), showing an association between coffee consumption greater than 2 cups/day (i.e. about 200mg) with amyloid PIB-PET positivity. Another recent study involving only cognitively normal subjects (mean age 71 years) also found a significant association between higher caffeine consumption and slower accumulation of cerebral amyloid load measured by the PIB-PET during a 126-month follow-up [32].

Several hypotheses might explain the effect of caffeine on amyloid pathology. Firstly, direct effects on amyloid peptide production have been described. Indeed, long-term administration of caffeine to mice overexpressing the amyloid peptide precursor protein (APP) has been associated with a reduction of Aβ levels in the cortex and hippocampus, as well as in brain interstitial fluid, probably by reducing the expression of γ-secretase and the APP cleavage enzyme (BACE1), the key enzymes involved in the production of Aβ from APP [29,55]. This would be in line with the ability of the A_2A_ receptor, an important pharmacological target of caffeine, to modulate Aβ production *in vitro* [56] and *in vivo* [57]. The effect of caffeine on the amyloid load would also rely on its ability to restrain neuroinflammatory processes which are known to impair the clearance of amyloid lesions by glial cells (see [58,59] for reviews). Non-mutually exclusively, caffeine might also act indirectly on the metabolism of amyloid peptide by increasing the production of CSF [60], leading to an increase in CSF turnover and, subsequently, to an improvement of Aβ clearance from the brain (for review, [61]). This attractive hypothesis remains however to be firmly demonstrated.

Overall, the strength of the present study relies on the possibility to address the impact of habitual caffeine consumption in a cohort solely including MCI and AD patients very well described at the clinical, neuropsychological, MRI and CSF biomarker levels; and the possibility to adjust the statistical analysis on known confounding factors for AD (APOEε4, age, sex, education level) and caffeine intake (smoking). The present study has, however, some limitations. Although the sample size is larger than in the previous study of Travasos et al. [35], it remains limited, in particular regarding the naMCI group. Other limitation of our cross-sectional study is that the associations observed do not allow us to establish with certainty a cause-and-effect relationship between caffeine consumption and the effects observed. Causality can only be demonstrated by a randomized double-blind interventional trial comparing patient treated with caffeine arm or a placebo. This is the reason why we have set up the interventional CAFCA clinical phase 3 trial (NCT04570085) which is currently recruiting.

## Data Availability

All data produced in the present study are available upon reasonable request to the authors

## CONFLICT OF INTEREST STATEMENT

David Blum DB is a (non-appointed) member of the scientific advisory board of Marvel Biosciences Corp developing an A_2A_R antagonist but has no conflict of interest regarding the present work. Olivier Hanon received personal payment from Bayer, Servier, AstraZeneca, Boston Scientific, Vifor, BMS, Boehringer-Ingelheim, and Pfizer for lectures and/or consulting services. Jean Sebastien Vidal received payment from Bayer for lectures made to non-profit medical association. Sylvain Lehmann received for his institution support from the following: H2020 MARIE SKŁODOWSKA-CURIE “MIRIADE Multi-omics Interdisciplinary Research Integration to Address DEmentia diagnosis,” ANR Flash Covid: “ProteoCOVID: Clinical proteomic characterization of the SARS-CoV-2 Spike protein to optimize its detection and the development of serological assays,” ANR “Silk_road: The Stable Isotope Labeling Kinetics (SILK) road to investigate human protein turnover in blood and cerebrospinal fluid,” EUROMET EMPIR “NeuroMet2 project: Metrology and Innovation for early diagnosis and accurate stratification of patients with neurodegenerative diseases.” During the past 36 months, he had a patent issued for “Procédé de préparation d’un échantillon peptidique” Brevet INPI n◦1905247 du 20/05/2019 du CHU DE MONTPELLIER, UNIVERSITÉ DE MONTPELLIER and SPOT TO LAB. He received personal payment for participating on the Roche Diagnostic board on CSF biomarkers. Stéphanie Bombois, Bernadette Allinquant, Christiane Baret-Rose, Jean-Marc Tréluyer, Hendy Abdoul, Patrick Gelé, Christine Delmaire, Jean-François Mangin, and Evelyne Galbrun have no conflicts of interest. Fredéric Blanc received honoraria from Roche and Biogen for presentations. He received payment to his institution as the national coordinator for the clinical trial DELPHIA for patients with dementia with Lewy bodies (Eisai). He received payment to his institution as the national coordinator for the clinical trial GRADUATE for patients with Alzheimer’s disease. Luc Buée received support for the present manuscript from LabEx DISTALZ. He received grants or contracts from the French National Research Agency (ANR) Fondation pour la Recherche Médicale (FRM). In the past 36 months, he had a patents on anti-tau therapy issued. Jacques Touchon received payment or honoraria as chairman of CTAD. He received contracts from Regenlife and consulting fees from Regenlife. He is Chairman of JT Conseil society. Jacques Hugon received grants or contracts from Protekt therapeutics, consulting fees from Protekt therapeutics. He is principal investigator of RECAGE project European Union H20/20 programme and he is member of the scientific board of Fondation Philippe Chatrier, Paris, France. Bruno Vellas received grants or contracts from Biogen, Roche, and Lilly; consulting fees from Roche, Lily, Biogen, and Cerellis; and is part of WHO’s ICOPE program (unpaid position). AthanBase Benetos is the president of the European Geriatric Medicine Society (unpaid position). He received support for attending meetings and/or travel from Fukuda company, for the Congress of the European Society of Hypertension, and received royalties or licenses from Cambridge University Editions. Gilles Berrut received a grant from Boehringer Ingelheim and consulting fees from Boehringer Ingelheim, Smart macadam Institut, bien vieillir Korian. Elena Paillaud has no conflicts of interest. David Wallon, Giovanni Castelnovo, Lisette Volpe-Gillot, Marc Paccalin, Philippe Robert, and Vincent Camus have no conflicts of interest. Olivier Godefroy received support to his institution for attending meetings and/or travel from BRISTOL-MYERS SQUIBB, ROCHE SAS, BIOGEN FRANCE SAS. Joël Belmin received consulting fees from Pfizer and honoraria from Novartis Pharma. Pierre Vandel is the président of the “Société Francophone de Psychogériatrie et Psychiatrie de la Personne Agée” (SF3PA) and received consulting fees from Eisai. Jean-Luc Novella, Emmanuelle Duron, Anne-Sophie Rigaud, Susanna Schraen-Maschke, Alain Duhamel, Nassima Ramdane, Lucie Vaudran, Caroline Dussart, Bernard Sablonnière and Audrey Gabelle have no conflicts of interest.

## FUNDING

The BALTAZAR study is supported by two grants from the French ministry of Health (Programme Hospitalier de Recherche Clinique, PHRC): PHRC2009/01-04 and PHRC-13-0404 and by the Foundation Plan Alzheimer. DB is supported by ANR JANUS (ANR-21-CE14-0053) and PHRC-I CAFCA.

## CONSENT STATEMENT

All participants or their legal guardians gave written informed consent.

## COLLABORATORS: The BALTAZAR study group

Olivier Hanon [1], Frédéric Blanc [2], Yasmina Boudali [1], Audrey Gabelle [3], Jacques Touchon [3], Marie-Laure Seux [1], Hermine Lenoir [1], Catherine Bayle [1], Stéphanie Bombois [4], Christine Delmaire [4], Xavier Delbeuck [5], Florence Moulin [1], Emmanuelle Duron [6], Florence Latour [7], Matthieu Plichart [1], Sophie Pichierri [8], Galdric Orvoën [1], Evelyne Galbrun [9], Giovanni Castelnovo [10], Lisette Volpe-Gillot [11], Florien Labourée [1], Pascaline Cassagnaud [12], Claire Paquet [13], Françoise Lala [14], Bruno Vellas [14], Julien Dumurgier [13], Anne-Sophie Rigaud [1], Christine Perret-Guillaume [15], Eliana Alonso [16], Foucaud du Boisgueheneuc [17], Laurence Hugonot-Diener [1], Adeline Rollin-Sillaire [12], Olivier Martinaud [18], Clémence Boully [1], Yann Spivac [19], Agnès Devendeville [20], Joël Belmin [21], Philippe Robert [22], Thierry Dantoine [23], Laure Caillard [1], David Wallon [24], Didier Hannequin [18], Nathalie Sastre [14], Sophie Haffen [25], Anna Kearney-Schwartz [15], Jean-Luc Novella [26], Vincent Deramecourt [12], Valérie Chauvire [27], Gabiel Abitbol [1], Nathalie Schwald [19], Caroline Hommet [28], François Sellal [29], Marie-Ange Cariot [16], Mohamed Abdellaoui [30], Sarah Benisty [31], Salim Gherabli [1], Pierre Anthony [29], Frédéric Bloch [32], Nathalie Charasz [1], Sophie Chauvelier [1], Jean-Yves Gaubert [1], Guillaume Sacco [22], Olivier Guerin [22], Jacques Boddaert [33], Marc Paccalin [17], Marie-Anne Mackowiak [12], Marie-Thérèse Rabus [9], Valérie Gissot [34], Athanase Benetos [15], Candice Picard [20], Céline Guillemaud [35], Gilles Berrut [8], Claire Gervais [22], Jacques Hugon [13], Jean-Marc Michel [29], JeanPhilippe David [19], Marion Paulin [12], Pierre-Jean Ousset [14], Pierre Vandel [36], Sylvie Pariel [21], Vincent Camus [37], Anne Chawakilian [1], Léna Kermanac’h [1], Anne-Cécile Troussiere [12], Cécile Adam [23], Diane Dupuy [20], Elena Paillaud [16], Hélène Briault [9], Isabelle Saulnier [38], Karl Mondon [37], Marie-Agnès Picat [23], Marie Laurent [16], Olivier Godefroy [20], Rezki Daheb [16], Stéphanie Libercier [29], Djamila Krabchi [1], Marie Chupin [39], JeanSébastien Vidal [1], Edouard Chaussade [1], Christiane Baret-Rose [40], Sylvain Lehmann [41], Bernadette Allinquant [40], Susanna Schraen-Maschke [4].

[1] Université de Paris, EA 4468, APHP, Hopital Broca, Memory Resource and Research Centre of de Paris-Broca-Ile de France, F-75013 Paris, France.
[2] Université de Strasbourg, Hôpitaux Universitaires de Strasbourg, CM2R, pôle de Gériatrie, Laboratoire ICube, FMTS, CNRS, équipe IMIS, F-67000 Strasbourg, France.
[3] Université de Montpellier, CHU Montpellier, Memory Research and Resources center of Montpellier, department of Neurology, Inserm INM NeuroPEPs team, excellence center of neurodegenerative disorders, F-34000 Montpellier, France.
[4] Univ. Lille, Inserm, CHU Lille, U1172-LilNCog, LiCEND, LabEx DISTALZ, F-59000 Lille, France.
[5] Univ. Lille, Inserm U1171 Degenerative and Vascular Cognitive Disorders, F-59000 Lille, France.
[6] Université Paris-Saclay, APHP, Hôpital Paul Brousse, département de gériatrie, Équipe MOODS, Inserm 1178, F-94800 Villejuif, France.
[7] Centre Hospitalier de la Côte Basque, Department of Gerontology, F-64100 Bayonne, France.
[8] Université de Nantes, EA 4334 Movement-Interactions-Performance, CHU Nantes, Memory Research Resource Center of Nantes, Department of clinical gerontology, F-44000 Nantes, France.
[9] Sorbonne Université, APHP, Centre Hospitalier Dupuytren, Department of Gérontology 2, F-91210 Draveil, France.
[10] CHU de Nimes, Hôpital Caremeau, Neurology Department, F-30029 Nimes, France.
[11] Hôpital Léopold Bellan, Service de Neuro-Psycho-Gériatrie, Memory Clinic, F-75014 Paris, France.
[12] Univ. Lille, CHU de Lille, Memory Resource and Research Centre of Lille, Department of Neurology, F-59000 Lille, France.
[13] Université de Paris, APHP, Groupe Hospitalier Saint Louis-LariboisièreFernand Widal, Center of Cognitive Neurology, F-75010 Paris, France.
[14] Université de Toulouse III, CHU La Grave-Casselardit, Memory Resource and Research Centre of Midi-Pyrénées, F-31300 Toulouse, France.
[15] Université de Lorraine, CHRU de Nancy, Memory Resource and Research Centre of Lorraine, F-54500 Vandoeuvre-lès-Nancy, France.
[16] Université de Paris, APHP, Hôpital europeen Georges Pompidou, Service de Gériatrie, F-75015, Paris, France.
[17] CHU de Poitiers, Memory Resource and Research Centre of Poitiers, F-86000 Poitiers, France.
[18] CHU Charles Nicolle, Memory Resource and Research Centre of HauteNormandie, F-76000 Rouen, France.
[19] APHP, Centre Hospitalier Émile-Roux, Department of Gérontology 1, F-94450 Limeil-Brévannes, France.
[20] CHU d’Amiens-Picardie, Memory Resource and Research Centre of AmiensPicardie, F-80000 Amiens, France.
[21] Sorbonne Université, APHP, Hôpitaux Universitaires Pitie-Salpêtrière-Charles-Foix, Service de Gériatrie Ambulatoire, F-75013 Paris, France.
[22] Université Côte d’Azur, CHU de Nice, Memory Research Resource Center of Nice, CoBTek lab, F-06100 Nice, France.
[23] CHU de Limoges, Memory Research Resource Center of Limoges, F-87000 Limoges, France.
[24] Normandie Univ, UNIROUEN, Inserm U1245, CHU de Rouen, Department of Neurology and CNR-MAJ, Normandy Center for Genomic and Personalized Medicine, CIC-CRB1404, F-76000, Rouen, France.
[25] CHU de Besançon, Memory Resource and Research Centre of Besançon Franche-Comté, F-25000 Besançon, France.
[26] Université de Reims Champagne-Ardenne, EA 3797, CHU de Reims, Memory Resource and Research Centre of Champagne-Ardenne, F-51100 Reims, France.
[27] CHU d’Angers, Memory Resource and Research Centre of Angers, F-49000 Angers, France.
[28] CHRU de Tours, Memory Resource and Research Centre of Tours, F-37000 Tours, France.
[29] Université de Strasbourg, CHRU de Strasbourg, Memory Resource and Research Centre of Strasbourg/Colmar, Inserm U-118, F-67000 Strasbourg, France.
[30] Univ Paris Est Creteil, EA 4391 Excitabilité Nerveuse et Thérapeutique, CHU Henri Mondor, Department of Neurology, F-94000 Créteil, France.
[31] Hôpital Fondation Rothschild, Department of Neurology, F-75019 Paris, France.
[32] CHU d’Amiens-Picardie, Department of Gerontology, F-80000 Amiens, France.
[33] Sorbonne Université, APHP, Hôpitaux Universitaires Pitie-Salpêtrière-Charles Foix, Memory Resource and Research Centre, Centre des Maladies Cognitives et Comportementales IM2A, Inserm UMR 8256, F-75013 Paris, France.
[34] Université François-Rabelais de Tours, CHRU de Tours, Memory Resource and Research Centre of Tours, Inserm CIC 1415, F-37000 Tours, France.
[35] Sorbonne Université, APHP, Hôpitaux Universitaires Pitie-Salpêtrière-Charles Foix, Memory Resource and Research Centre, Centre des Maladies Cognitives et Comportementales IM2A, F-75013 Paris, France.
[36] Université Bourgogne Franche-Comté, Laboratoire de Recherches Intégratives en Neurosciences et Psychologie Cognitive, CHU de Besançon, Memory Resource and Research Centre of Besançon Franche-Comté, F-25000 Besançon, France.
[37] Université François-Rabelais de Tours, CHRU de Tours, UMR Inserm U1253, F-37000 Tours, France.
[38] Université de Limoges, EA 6310 HAVAE, CHU de Limoges, Memory Research Resource Center of Limoges, F-87000 Limoges, France.
[39] Université Paris-Saclay, Neurospin, CEA, CNRS, cati-neuroimaging.com, CATI Multicenter Neuroimaging Platform, F-91190 Gif-sur-Yvette, France.
[40] Université de Paris, Institute of Psychiatric and Neurosciences, Inserm UMR-S 1266, F-75014 Paris, France.
[41] Université de Montpellier, CHU Montpellier, LBPC, Inserm, F-34000 Montpellier, France.

## Notes

### Competing Interest Statement

David Blum DB is a (non-appointed) member of the scientific advisory board of Marvel Biosciences Corp developing an A2AR antagonist but has no conflict of interest regarding the present work. Olivier Hanon received personal payment from Bayer, Servier, AstraZeneca, Boston Scientific, Vifor, BMS, Boehringer-Ingelheim, and Pfizer for lectures and/or consulting services. Jean Sebastien Vidal received payment from Bayer for lectures made to non-profit medical association. Sylvain Lehmann received for his institution support from the following: H2020 MARIE SKŁODOWSKA-CURIE MIRIADE Multi-omics Interdisciplinary Research Integration to Address DEmentia diagnosis, ANR Flash Covid: ProteoCOVID: Clinical proteomic characterization of the SARS-CoV-2 Spike protein to optimize its detection and the development of serological assays, ANR Silk_road: The Stable Isotope Labeling Kinetics (SILK) road to investigate human protein turnover in blood and cerebrospinal fluid, EUROMET EMPIR NeuroMet2 project: Metrology and Innovation for early diagnosis and accurate stratification of patients with neurodegenerative diseases. During the past 36 months, he had a patent issued for Procede de preparation d un echantillon peptidique Brevet INPI n◦1905247 du 20/05/2019 du CHU DE MONTPELLIER, UNIVERSITE DE MONTPELLIER and SPOT TO LAB. He received personal payment for participating on the Roche Diagnostic board on CSF biomarkers. Stephanie Bombois, Bernadette Allinquant, Christiane Baret-Rose, Jean-Marc Treluyer, Hendy Abdoul, Patrick Gele, Christine Delmaire, Jean-Francois Mangin, and Evelyne Galbrun have no conflicts of interest. Frederic Blanc received honoraria from Roche and Biogen for presentations. He received payment to his institution as the national coordinator for the clinical trial DELPHIA for patients with dementia with Lewy bodies (Eisai). He received payment to his institution as the national coordinator for the clinical trial GRADUATE for patients with Alzheimer s disease. Luc Buee received support for the present manuscript from LabEx DISTALZ. He received grants or contracts from the French National Research Agency (ANR) Fondation pour la Recherche Medicale (FRM). In the past 36 months, he had a patent on anti-tau therapy issued. Jacques Touchon received payment or honoraria as chairman of CTAD. He received contracts from Regenlife and consulting fees from Regenlife. He is Chairman of JT Conseil society. Jacques Hugon received grants or contracts from Protekt therapeutics, consulting fees from Protekt therapeutics. He is principal investigator of RECAGE project European Union H20/20 programme and he is member of the scientific board of Fondation Philippe Chatrier, Paris, France. Bruno Vellas received grants or contracts from Biogen, Roche, and Lilly; consulting fees from Roche, Lily, Biogen, and Cerellis; and is part of WHO s ICOPE program (unpaid position). AthanBase Benetos is the president of the European Geriatric Medicine Society (unpaid position). He received support for attending meetings and/or travel from Fukuda company, for the Congress of the European Society of Hypertension, and received royalties or licenses from Cambridge University Editions. Gilles Berrut received a grant from Boehringer Ingelheim and consulting fees from Boehringer Ingelheim, Smart macadam Institut, bien vieillir Korian. Elena Paillaud has no conflicts of interest. David Wallon, Giovanni Castelnovo, Lisette Volpe-Gillot, Marc Paccalin, Philippe Robert, and Vincent Camus have no conflicts of interest. Olivier Godefroy received support to his institution for attending meetings and/or travel from BRISTOL-MYERS SQUIBB, ROCHE SAS, BIOGEN FRANCE SAS. Joel Belmin received consulting fees from Pfizer and honoraria from Novartis Pharma. Pierre Vandel is the president of the Societe Francophone de Psychogeriatrie et Psychiatrie de la Personne Agee (SF3PA) and received consulting fees from Eisai. Jean-Luc Novella, Emmanuelle Duron, Anne-Sophie Rigaud, Susanna Schraen-Maschke, Alain Duhamel, Nassima Ramdane, Lucie Vaudran, Caroline Dussart, Bernard Sablonniere and Audrey Gabelle have no conflicts of interest.

### Clinical Protocols

https://classic.clinicaltrials.gov/ct2/show/NCT01315639

### Author Declarations

This study is ancillary to BALTAZAR (Biomarker of AmyLoid pepTide and AlZheimer s diseAse Risk), a multicenter (23 memory centers) prospective cohort study (ClinicalTrials.gov Identifier #NCT01315639) including participants with mild cognitive impairments (MCI) and Alzheimer s Disease (AD) at baseline from September 2010 to April 2015 and with an ongoing 3-year follow-up. All participants or their legal guardians gave written informed consent. The study was approved by the Paris Ethics Committee (CPP Ile de France IV Saint Louis Hospital).

